# Development and comparison of a novel multiple cross displacement amplification (MCDA) assay with other nucleic acid amplification methods for SARS-CoV-2 detection

**DOI:** 10.1101/2020.10.03.20206193

**Authors:** Laurence Don Wai Luu, Michael Payne, Xiaomei Zhang, Lijuan Luo, Ruiting Lan

## Abstract

The development of alternative isothermal amplification assays including multiple cross displacement amplification (MCDA) may address speed and portability limitations of real-time PCR (rt-PCR) methods for SARS-CoV-2 detection. We developed a novel SARS-CoV-2 MCDA assay and compared its speed and sensitivity to loop-mediated isothermal amplification (LAMP) and rt-PCR. Two MCDA assays targeting SARS-CoV-2 N gene and ORF1ab was designed. The fastest time to detection and sensitivity of MCDA was compared to LAMP and rt-PCR using DNA standards and transcribed RNA. For N gene, MCDA was faster than LAMP and rt-PCR by 10 and 20 minutes, respectively with fastest time to detection at 5.2 minutes. rt-PCR had highest sensitivity with limit of detection at 10 copies/µl compared with MCDA (100 copies/µl) and LAMP (500 copies/µl). For ORF1ab, MCDA and LAMP had similar speed with fastest time to detection at 9.7 and 8.4 minutes, respectively. LAMP was more sensitive for ORF1ab detection with 50 copies/µl compared to MCDA (500 copies/µl). In conclusion, different nucleic acid amplification methods provide different advantages. MCDA is the fastest nucleic acid amplification method for SARS-CoV-2 while rt-PCR is the most sensitive. These advantages should be considered when determining the most suitable nucleic acid amplification methods for different applications.

## Introduction

Rapid, portable and highly sensitive assays are essential to controlling the COVID-19 pandemic. Real-time-PCR (rt-PCR) is the gold standard for detection of SARS-CoV-2 genetic material^1^. However, rt-PCR requires trained personnel, advanced equipment and relatively long assay times making it unsuitable for large-scale community screening. Other tests developed include serological assays that rely on IgM/IgG antibodies which takes ∼5 days to appear after symptom onset making them unsuitable for rapid early detection^2^.

The development of alternative nucleic acid amplification methods including loop-mediated isothermal amplification (LAMP) may offer improved speed, sensitivity and portability for SARS-CoV-2 detection^3^. Another isothermal nucleic acid amplification method, called multiple cross displacement amplification (MCDA) which uses 10 primers instead of six, has also been suggested to have even higher sensitivity and speed than LAMP but has not yet been developed for SARS-CoV-2 detection^4,5^.

Despite claims of increased speed and sensitivity from isothermal amplification methods, no study has directly compared the speed and sensitivity of these three different nucleic acid amplification methods. Hence, here we developed an MCDA assay for SARS-CoV-2 detection and compared its speed and sensitivity to existing LAMP and rt-PCR methods.

## Methods

### MCDA target gene selection

To identify target genes with highly conserved regions and a suitable GC-content for MCDA, 1,216 SARS-CoV-2 genomes deposited in GISAID (all available complete, high coverage genomes (>29,000 bp) with low coverage flags excluded up until March 26, 2020)^6^ were aligned against the SARS-CoV-2 reference genome: NC_045512.2 using Snippy (v4.3.6) with the -ctgs flag and default settings (https://github.com/tseemann/snippy). A sliding window approach was then applied to identify conserved 300bp windows with GC content >43%, low SNP number, and low total SNP number (total SNPs was defined as the number of strains with a SNP in a given window). Three 300 bp conserved regions were identified and selected for MCDA primer design with two regions in ORF1ab (NC_045512.2: 515-831 and 12968-13288) and one in the *N* gene (NC_045512.2: 28345-28647).

### MCDA primer design

For each region, 4 sets of MCDA primers were designed as previously described^5^. Each primer set consisted of 2 cross-primers (CP1/CP2), 2 displacement primers (F1/F2) and 6 amplification primers (C1/C2, D1/D2, R1/R2) (Supplementary Table 1). Non-specific primer binding was assessed using BLASTN against 14 non-SARS-CoV-2 coronaviruses used in Lamb *et al*.^7^, human genome (hg19) and 11 other common bacterial and viral respiratory pathogen/microbiome species.

**Table 1:** Comparison of the sensitivity and time to detection for MCDA, LAMP and rt-PCR targeting the N gene from 3 independent runs.

| DNA copies<br>per µl | MCDA average<br>detection time (min) | MCDA<br>reps<sup>\$</sup> | LAMP average<br>detection time (min) | LAMP reps<sup>\$</sup> | rt-PCR average Ct<br>value | rt-PCR average<br>detection time (min) | rt-PCR rep<sup>\$</sup> |
| --- | --- | --- | --- | --- | --- | --- | --- |
| 1 | NA | NA | NA | NA | 43.93±1.34 | 38.61±1.11 | 3/9 |
| 10 | 19.3 ±17.6 | 5/9 | 20.9 ±0.4 | 2/9 | **35.5 ±0.6\*** | **31.6 ±0.47\*** | **9/9\*** |
| 25 | 20.4 ±22.4 | 4/9 | 26.7 ±11.4 | 3/9 | 34.2 ±0.03 | 30.5 ±0.03 | 9/9 |
| 50 | 7.8 ±1 | 6/9 | 20.3 ±1 | 6/9 | 33.0 ±0.25 | 29.5 ±0.21 | 9/9 |
| 100 | **10.2 ±3.2\*** | **9/9\*** | 23.5 ±8.6 | 5/9 | 32.4 ±0.22 | 29.0 ±0.19 | 9/9 |
| 5,00 | 6.5 ±0.1 | 9/9 | **17.5 ±0.6\*** | **9/9\*** | 30.3 ±0.68 | 27.3 ±0.57 | 9/9 |
| 1,000 | 6.2 ±0.2 | 9/9 | 17.3 ±0.7 | 9/9 | 29.2 ±0.26 | 26.3 ±0.22 | 9/9 |
| 10,000 | 5.2 ±0.1 | 9/9 | 15.0 ±0.4 | 9/9 | 25.7 ±0.06 | 23.4 ±0.05 | 9/9 |

NA = no amplification detected
\*Limit of detection
<sup>\$</sup> reps: 3 runs with 3 technical replicates = 9 replicates.

### Preparation of DNA/RNA standards

For each region, ∼500 bp gene fragments for ORF1ab (NC_045512.2: 416-931 and 12869-13388) and N gene (NC_045512.2: 28246-28747) were synthesised with an additional 100 bp up and downstream of the target region (ThermoFisher) (Supplementary Table 2). Each fragment contained a T7 promoter for transcription and M13 adapters for amplification.

**Table 2:**
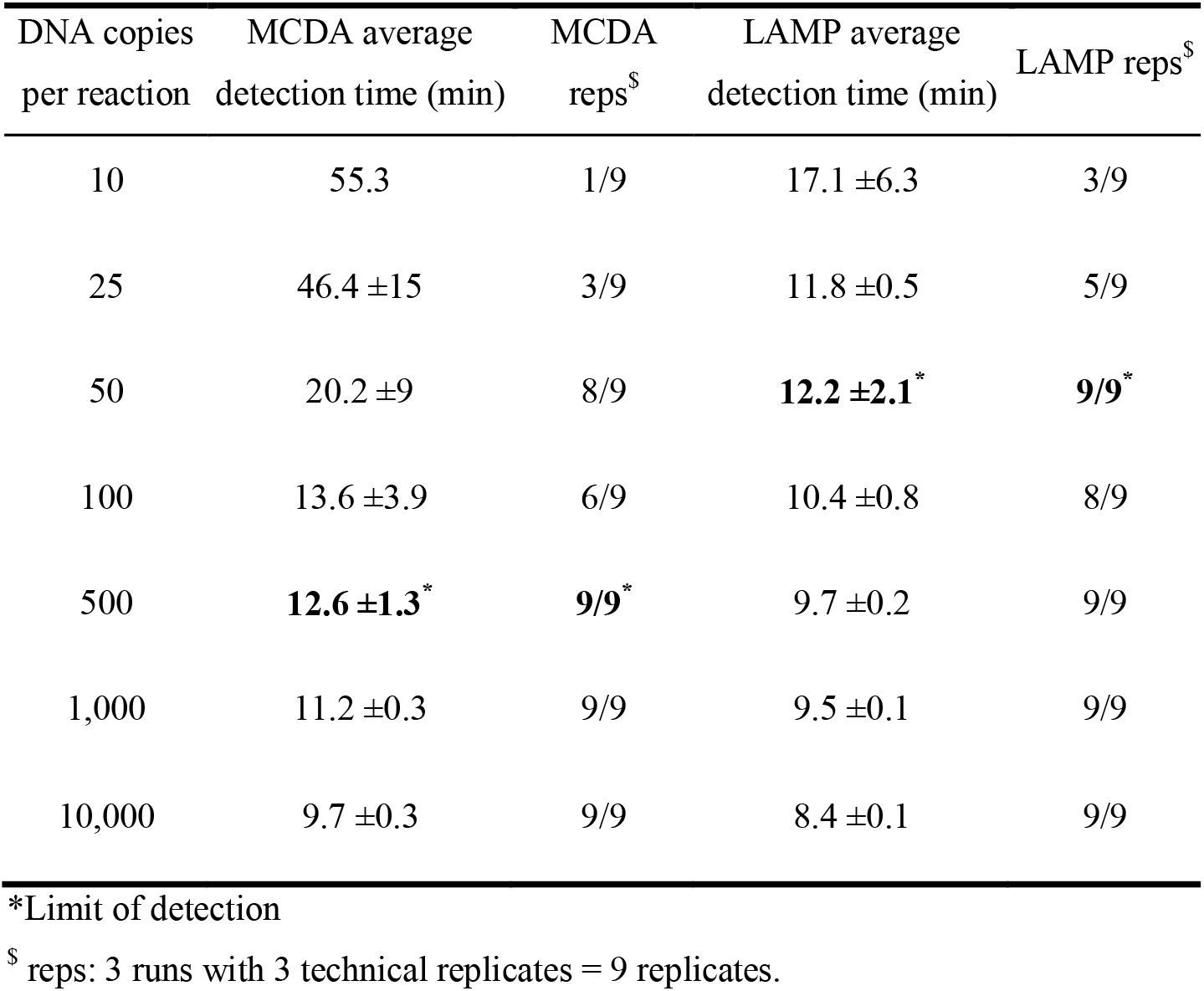
Comparison of the sensitivity and time to detection for MCDA and LAMP targeting ORF1ab from 3 independent runs.

Seven DNA standards from synthesised gene fragments were prepared for ORF1ab (100,000, 10,000, 5000, 1000, 500, 250 and 100 copies/µl) while eight DNA standards were prepared for the N gene (100,000, 10,000, 5000, 1000, 500, 250, 100 and 10 copies/µl).

For RNA, 1 pg of synthesised gene fragments were transcribed overnight at 37°C using T7 RNA polymerase (Sigma). Overnight DNA digestion was performed using the turbo DNA free kit (ThermoFisher) and further treated with DNase I (NEB) until all traces of DNA were removed. Complete DNA removal was confirmed after each round of DNase treatment using rt-PCR with the SensiFAST SYBR kit (Bioline) and F1/R1 MCDA primers (Supplementary Table 1). The transcribed RNA was serially diluted and used as input. Since the amount of RNA transcribed was below the 250 pg/µl limit of detection for qubit HS RNA assay (ThermoFisher), the input RNA copy number could not be determined. Therefore, the lowest detectable dilutions were used for sensitivity comparison.

### Initial evaluation of MCDA primer sets and optimisation of isothermal amplification temperature

MCDA reactions were performed using the WarmStart LAMP (DNA and RNA) kit (NEB) which contains a warmstart RTx reverse transcriptase and Bst2.0 polymerase for simultaneous reverse transcription and isothermal amplification. Antarctic thermolabile UDG was also added in each reaction to prevent carryover contamination.

For each primer set, a primer mix containing: 3.3 μM of F1 and F2, 6.67 μM of C1 and C2, 10 μM of R1, R2, D1 and D2 and 20 μM of CP1 and CP2 was used. Standard desalting purified primers were used for the initial evaluation and optimisation tests while HPLC purified primers were used for sensitivity and speed comparison against LAMP and rt-PCR tests.

For the initial evaluation of each MCDA primer set, a 10 µl reaction was used and contained: 5 µl of 2x WarmStart master mix, 0.2 µl of fluorescent dye, 1.2 µl of MCDA primer mix, 0.2 µl of 1U/µl Antarctic thermolabile UDG (NEB), 0.7 µl of 10 mM dUTP, 1.7 µl of H_2_O and 1 µl of 1000 copy/µl DNA template (final reaction concentration = 100 DNA copies/µl). The

final concentration of each MCDA primer in the reaction was 0.4 μM of F1 and F2, 0.8 μM of C1 and C2, 1.2 μM of R1, R2, D1 and D2 and 2.4 μM of CP1 and CP2. Specificity of MCDA primer sets were also evaluated using purified human genomic DNA (Sigma) and a microbial community DNA standard (Zymo Research). MCDA reactions were performed in triplicates in the Rotor-Gene Q (Qiagen) with isothermal amplification at either 60°C, 63°C or 65°C for 1 h and real time fluorescence detection every 60 seconds, followed by enzyme inactivation at 95°C for 5 min and a final melt curve from 50°C – 99°C to ensure correct MCDA product.

### Comparison of MCDA, LAMP and rt-PCR

To compare the speed and sensitivity (limit of detection) of MCDA, LAMP and rt-PCR, published primers targeting the same SARS-CoV-2 MCDA ORF1ab (NC_045512.2: 416-931) and N (NC_045512.2: 28246-28747) regions were used (Supplementary Table 1). For LAMP, two published primer sets from Zhang et al.^3^ which targeted the same region as our MCDA were compared. For rt-PCR, there were no suitable published primers pairs which targeted the same ORF1ab region, therefore only primers submitted by the National Institute of Health, Thailand against the N gene was compared^8^. All primers were HPLC-purified grade.

MCDA, LAMP and rt-PCR were tested in three independent runs (biological replicates) using the same aliquot of DNA/RNA. Each run contained 3 technical replicates. The limit of detection was defined as the highest dilution where all 9 replicates (3 biological replicates x 3 technical replicates) were detected.

To reduce between run variations, 10 µl MCDA and LAMP reactions were set up and performed simultaneously in the same run. MCDA reactions were prepared as described above. For LAMP, each 10 µl reaction contained: 5 µl 2x WarmStart master mix (NEB), 0.2 µl 50x fluorescent dye (NEB), 0.2 µl 1U/µl Antarctic thermolabile UDG (NEB), 0.7 µl 10 mM dUTP, 1 µl LAMP primer mix, 1.9 µl of H_2_O and 1 µl of DNA/RNA template. Each LAMP primer mix contained 16 µM FIP and BIP, 2 µM F3 and B3 and 4 µM LF and LB. The final concentration of each LAMP primer in the reaction was 1.6 µM FIP and BIP, 0.2 µM F3 and B3 and 0.4 µM LF and LB. MCDA and LAMP isothermal amplification was performed at 65°C as described above. The normalised fluorescence threshold line for N gene amplification was set above the background fluorescence at 0.2 for MCDA and LAMP. For ORF1ab MCDA and LAMP, the normalised fluorescence threshold line was set at 0.4 as background fluorescence was higher. The detection time for MCDA and LAMP was defined as the time it takes for the fluorescence intensity to pass the threshold line.

For rt-PCR using DNA templates, 10 µl reactions containing 5 µl SensiFAST probe No-ROX mix (Bioline), 0.5 µl rt-PCR primer mix (40 µM F and R, 10 µM probe), 3.5 µl of H_2_O and 1 µl DNA template were used. The final concentration of each rt-PCR primer and probe in the reaction was 2 µM F and R and 0.5 µM probe. The cycling conditions were 95°C for 2 min, followed by 45 cycles of 95°C for 15 secs and 55°C for 30 secs.

For rt-PCR with RNA templates, 10 µl reactions were set up containing 5 µl SensiFAST probe No-ROX One-Step mix (Bioline), 0.5 µl primer mix (40 µM F and R, 10 µM probe), 0.1 µl reverse transcriptase (Bioline), 3.4 µl of H_2_O and 1 µl RNA template. Reverse transcription was performed at 45°C for 20 minutes followed by rt-PCR amplification as described above for DNA.

To compare the speed of rt-PCR, cycle threshold (Ct) was converted to time using the following equation: Time = (Ct x 50 sec) + 120 sec. The detection time required for rt-PCR was calculated based on the cycling conditions (45 sec per cycle plus an initial 120 sec hold) and the ramp rate for the Rotor-gene Q (5 sec per cycle). The ramp rate for the Rotor gene Q is 15°C/s for heating and 20°C/s for cooling according to the manufacture’s technical information (https://www.qiagen.com/us/resources/download.aspx?id=2120af5e-8daf-4184-b277-aeb6ef5bbc05&lang=it-IT).

## Results

### Development of MCDA assays for SARS-CoV-2 detection

Three 300 bp conserved regions suitable for MCDA primer design were identified from the genome alignment of 1,216 SARS-CoV-2 strains. Two regions, designated as region 1 and 2, belonged to the ORF1ab gene at NC_045512.2 position 515-831 and 12968-13288 respectively. One region, designated as region 3, corresponded to the N-gene at NC_045512.2 position 28345-28647. Four MCDA primer sets for each region was designed and evaluated (Supplementary Table 1).

Each MCDA primer set was initially tested at 3 isothermal amplification temperatures (60°C, 63°C and 65°C) using 1000 DNA copies/reaction as the starting template. As seen in Figure 1, regardless of the primer sets used, the slowest amplification time was observed at 60°C. Amplification at 63°C and 65°C were similar and 65°C was chosen as the isothermal amplification temperature used.

**Figure 1:**
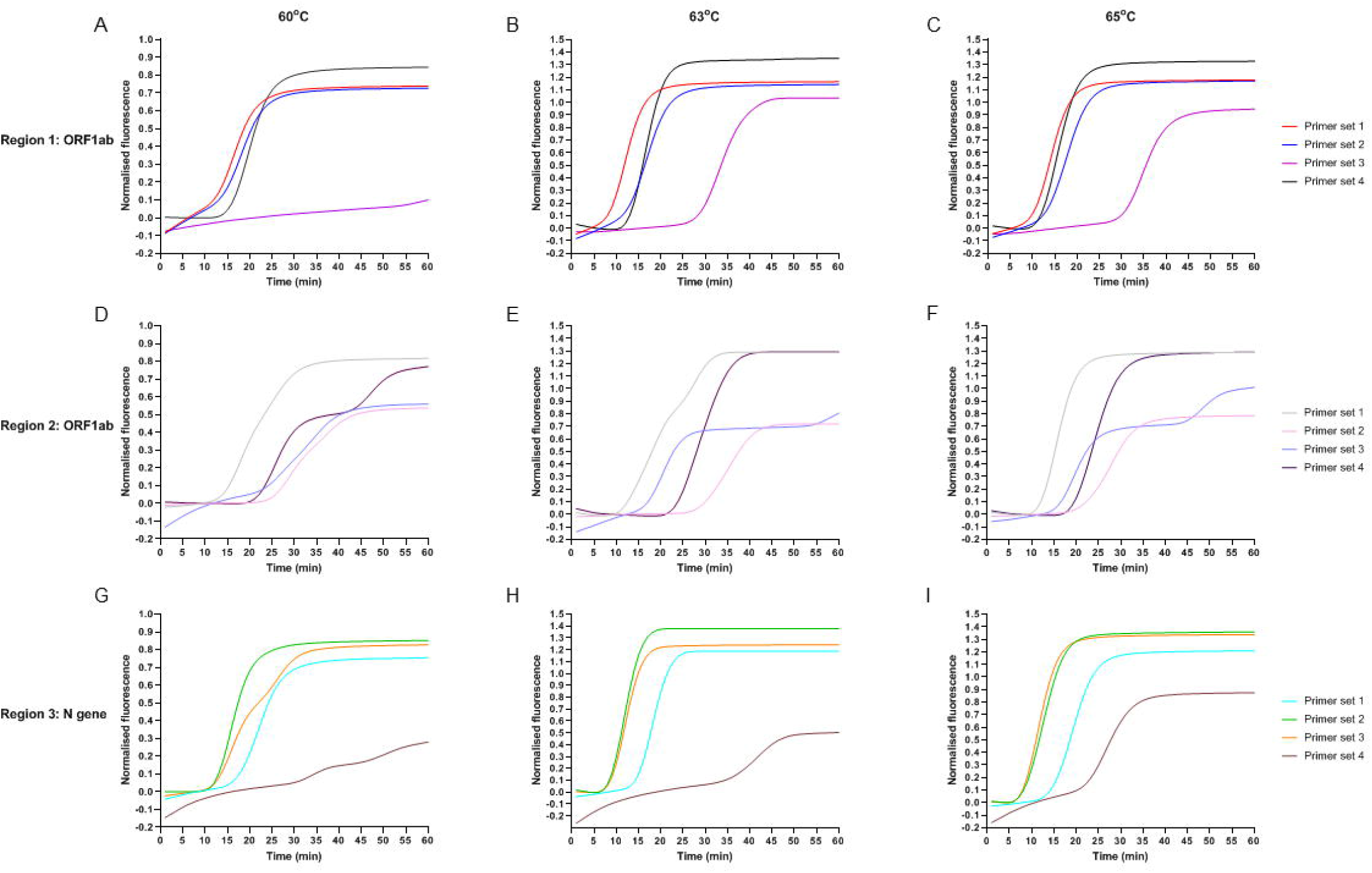
Initial evaluation of MCDA primer sets at 3 different isothermal amplification temperature (60°C, 63°C and 65°C). Four MCDA primer sets were designed for each target region chosen. **A-C**: Region 1 ORF1ab: 515-831 **D-E:** Region 2 ORF1ab: 12968-13288 **F-H**: Region 3 N gene:28345-28647.

To maintain MCDA assay robustness against SNPs which may affect MCDA primer binding and amplification efficiency, primer sets from two different regions were chosen for further development as a duplex assay. Amplification of region 2 was the slowest for all primer sets (Figure 1 D-F) compared with region 1 and 3, taking between 15-25 minutes at 65°C. Primer sets in region 2 also had very high variation between technical replicates (data not shown). Therefore region 2 was removed from further evaluation.

Region 3 amplification of the N gene was the fastest with primer set 2 followed closely by primer set 3 (Figure 1G-I). Primer set 1 and 4 were the slowest for region 3 and were therefore eliminated from further testing. We also observed that primer set 2 had tighter technical replicates compared to primer set 3 (data not shown), thus region 3 primer set 2 was chosen as our final MCDA primer set for further sensitivity and specificity testing.

Within region 1, primer set 3 was the slowest with fluorescence appearing at ∼35 min (Figure 1C). This primer set was removed from further consideration. Primer set 1 was the fastest primers to amplify region 1 and was chosen for inclusion in our MCDA assay.

Therefore, the final primer sets chosen for MCDA SARS-CoV-2 detection was region 1 (ORF1ab) primer set 1 and region 3 (N gene) primer set 2 (Figure 2). Both primer sets showed no non-specific amplification when tested against human and microbial community genomic DNA.

**Figure 2:**
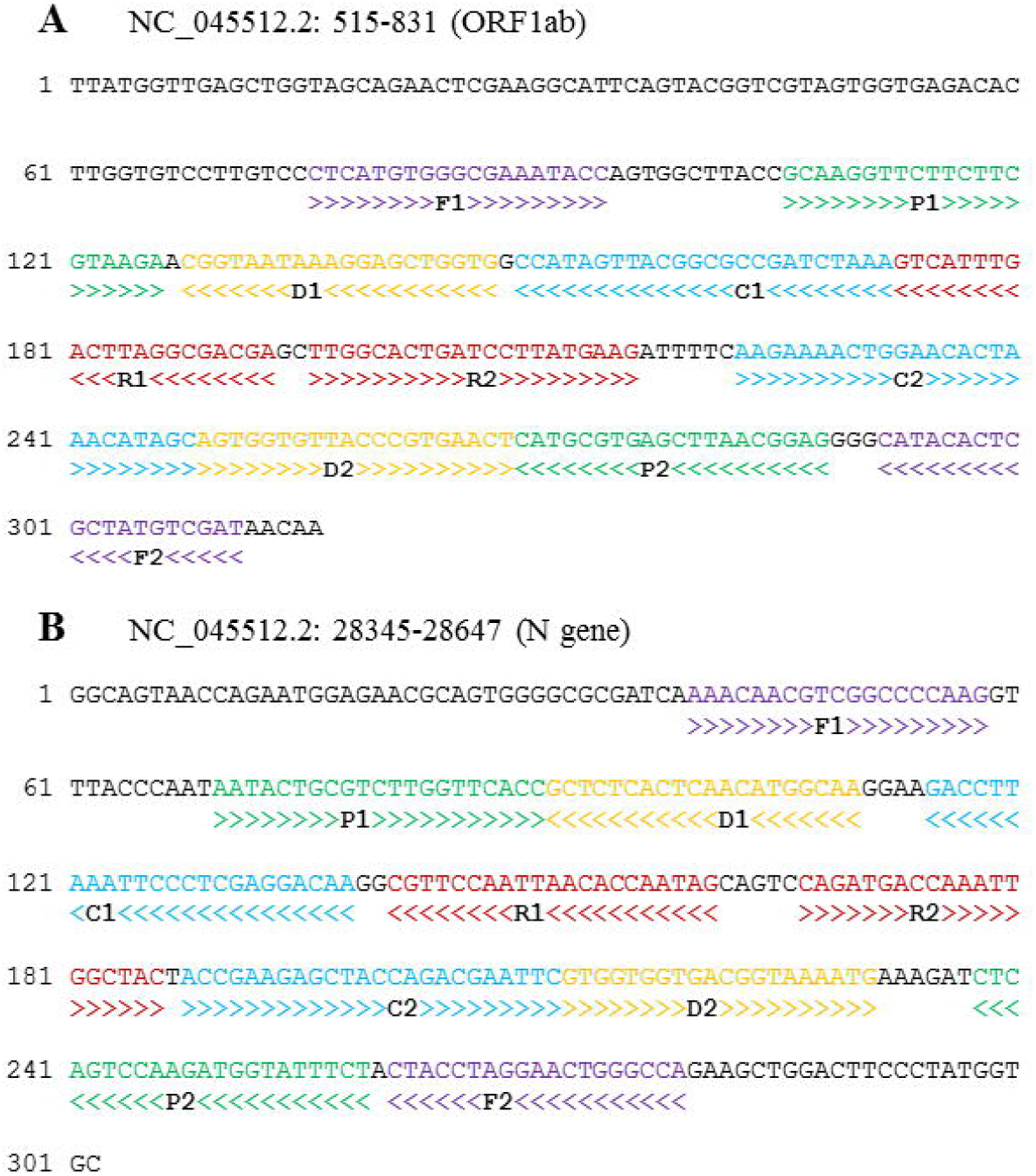
The nucleotide sequences and position of the final **(A)** ORF1ab and **(B)** N gene COVID-19 MCDA primer sets chosen in this study. Right and left arrows show sense and complementary sequences, respectively while coloured text indicate the position of primers: F1/F2 displacement primers in purple, P1/P2 primers in green, C1/C2 amplification primers in blue, D1/D2 amplification primers in yellow and R1/R2 amplification primers in red.

### Sensitivity and time to detection comparison of MCDA, LAMP and rt-PCR

The sensitivity and speed for MCDA, LAMP and rt-PCR were then compared for two SARS-CoV-2 genes. For the N gene (region 3), detection by MCDA was consistently faster than LAMP, by ∼10-13 minutes, for most DNA dilutions tested (Table 1). The average fastest detection time for MCDA was 5.2 minutes at 10,000 copies/µl while for LAMP it was 15 minutes. MCDA also had higher sensitivity with the limit of detection at 100 copies/µl while for LAMP it was 500 copies/µl. This limit of detection was equivalent to a rt-PCR Ct value of 32.4 and 30.3. respectively. A greater number of sporadic detections for higher dilutions were also observed for MCDA compared to LAMP. MCDA was also significantly faster than rt-PCR by ∼20 minutes for lower dilutions and 10 minutes for higher dilutions. At 10,000 copies/µl, the detection time for rt-PCR was 23 minutes. However, rt-PCR had the highest sensitivity with the limit of detection at 10 copies/µl and sporadic amplification at 1 copy/µl. Similar results were also observed using RNA template with rt-PCR being the most sensitive technique, detecting RNA at 10^−6^ dilution. For MCDA, the lowest RNA dilution detected was 10^-4^, with only sporadic detection for LAMP at this dilution.

For ORF1ab, LAMP was more sensitive than MCDA with the limit of detection at 50 copies/µl and had more sporadic detection at higher dilutions (Table 2). The limit of detection for MCDA was 500 copies/µl. LAMP was also slightly faster than MCDA by ∼1-3 minutes for concentrations above the limit of detection. The fastest time to detection for ORF1ab LAMP was 8.4 minutes while for MCDA it was 9.7 minutes.

## Discussion

Our results showed that MCDA is the fastest nucleic acid amplification method tested for SARS-CoV-2 detection with detection of the N gene as fast as 5 minutes. However, this was contingent on the gene targeted and the primer design with the NEB designed LAMP assay for ORF1ab ^3^ showing similar speed to our equivalent MCDA ORF1ab assay.

rt-PCR remains the most sensitive nucleic acid amplification method for SARS-CoV-2 detection compared to MCDA and LAMP. This result is in agreement with previous LAMP SARS-CoV-2 assays which showed rt-PCR having greater sensitivity^9,10^. The limit of detection for our MCDA N gene assay was 100 copies/µl or an equivalent N gene average Ct value of 32.4 (Table 1). The median rt-PCR Ct value in 324 clinical COVID-19 samples from a range disease severity was found to be 31.15 in Singanayagam et al.^11^ while in Passomsub et al.^12^ the median N gene Ct value in saliva samples and nasopharyngeal/throat samples were 31.8 and 30.5, respectively. This suggests that our MCDA assay has the potential to detect SARS-CoV-2 but with lower sensitivity and consistent with our comparison using synthetic templates. Furthermore, Lamb et al.^7^ developed a COVID-19 LAMP assay with a limit of detection of 0.08 fg or an equivalent rt-PCR Ct value of 30.3 and were able to validate their LAMP assay in 19/20 positive clinical COVID samples. Our MCDA assay has increased sensitivity and speed compared to LAMP, suggesting that MCDA has the potential for similar applications as LAMP with better sensitivity and speed.

For MCDA, this is the first study to directly benchmark the speed and sensitivity of MCDA to rt-PCR against the same targets. Previous MCDA studies only compared gel-based PCR^4^, different rt-PCR gene targets^13,14^ or used rt-PCR sensitivity results previously reported in other studies^4,5^ (as 100 copies in different studies may not be equivalent due to pipetting differences, differences in the method used to measure nucleic acid concentration (nanodrop vs qubit) or differences in machine calibration, etc.). In order to benchmark different nucleic acid techniques, we used and recommend the same reaction volume, same machine, same DNA standards and aliquots, and where possible the same run is used.

This study found that different nucleic acid amplification methods offer different advantages and this should be considered depending on the application. rt-PCR was the most sensitive method tested and should remain the gold standard for SARS-CoV-2 detection. However, the portable nature and speed of MCDA makes it suitable for settings where rt-PCR would be too slow. Although the fastest time to detection for MCDA is ∼5 minutes, MCDA amplification should be performed for at least 20 minutes to ensure reliable results for negative samples while for rt-PCR, the current amplification time, not including reverse transcription, is 30-40 minutes. Additionally, reverse transcription and amplification for MCDA and LAMP can occur simultaneously. This removes the need to sequentially perform an initial 20 min reverse transcription step prior to amplification as required for rt-PCR, making MCDA even faster. Therefore, it is estimated that the total time saved using MCDA compared to rt-PCR is 30-40 minutes. An additional advantage of MCDA is that it uses the same Bst polymerase and reverse transcriptase as LAMP, which are more resistant against inhibitors than rt-PCR^15^. LAMP has been shown to amplify SARS-CoV-2 RNA extracted using simple extraction procedures such as boiling^16-18^. Therefore, it is anticipated that MCDA can also be used to detect SARS-CoV-2 RNA extracted using these same procedures.

MCDA (and other isothermal amplification methods) is less sensitive than rt-PCR, making it less attractive to develop it further as a clinical diagnostic test. However, there may be situations where these methods will be useful such as rapid screening of samples with high viral RNA content. The addition of a colorimetric dye instead of a fluorescent dye can further simplify MCDA for rapid screening. Further studies in a variety of settings will be required to determine where MCDA and other isothermal nucleic acid methods can offer an advantage in certain settings where rapid test turnaround time or test simplicity is paramount.

## Data Availability

All data is provided in the manuscript and/or in the supplementary materials.

## Competing interests statement

The authors declare no competing interests.

## Author Contributions

RL conceived the study. LDWL performed the experiments, analysed the results and drafted the manuscript. MP performed the MCDA target selection and BLAST. XZ and LL designed the MCDA primers. All authors provided critical revision of the manuscript.

## Funding

This work was supported by a UNSW school research grant.

## Data availability

All data generated or analysed during this study are included in this published article (and its Supplementary Information files).

## Supplementary Materials

**Supplementary Table 1:** List of MCDA, LAMP and rt-PCR primers used in this study. Bolded MCDA primer names are primers shared between 2 or more primer sets. * indicate primers used in rt-PCR for confirmation of complete DNA removal from transcribed RNA. Red text indicates the final primer sets chosen for the MCDA SARS-CoV-2 assay.

**Supplementary Table 2:** List of synthesised gene fragments used as DNA/RNA template for MCDA, LAMP and rt-PCR. Blue indicates universal M13 adapters while red depicts the sequence for T7 promoter.

## Notes

### Competing Interest Statement

The authors have declared no competing interest.

### Author Declarations

This research does not include clinical samples therefore no oversight approval was required.

