## Supplementary Table 1 for "Development and comparison of a novel multiple cross displacement amplification (MCDA) assay with other nucleic acid amplification methods for SARS-CoV-2 detection"

**Supplementary Table 1:** List of MCDA, LAMP and rt-PCR primers used in this study. Bolded MCDA primer names are primers shared between 2 or more primer sets. * indicate primers used in rt-PCR for confirmation of complete DNA removal from transcribed RNA. Red text indicates the final primer sets chosen for the MCDA SARS-CoV-2 assay.

| MCDA Primer name | Sequence 5' --> 3' | Reference |
|---|---|---|
| **Region 1 ORF1ab primer set 1** | | |
| R1_1_F1* | CTCATGTGGGCGAAATACC | This study |
| **R1_1_2_F2** | ATCGACATAGCGAGTGTATG | |
| **R1_1_2_C1** | TTTAGATCGGCGCCGTAACTATGG | |
| **R1_1_2_C2** | AAGAAAACTGGAACACTAAACATAGC | |
| **R1_1_2_D1** | CACCAGCTCCTTTATTACCG | |
| R1_1_D2 | AGTGGTGTTACCCGTGAACT | |
| **R1_1_4_R1*** | TCGTCGCCTAAGTCAAATGAC | |
| R1_1_2_R2 | TTGGCACTGATCCTTATGAAG | |
| R1_1_2_CP1 | TTTAGATCGGCGCCGTAACTATGGGCAAGGTTCTTCTTCGTAAGA | |
| R1_1_CP2 | AAGAAAACTGGAACACTAAACATAGCCTCCGTTAAGCTCACGCATG | |
| **Region 1 ORF1ab primer set 2** | | |
| R1_2_F1 | ATGTGGGCGAAATACCAGTG | This study |
| **R1_1_2_F2** | ATCGACATAGCGAGTGTATG | |
| **R1_1_2_C1** | TTTAGATCGGCGCCGTAACTATGG | |
| **R1_1_2_C2** | AAGAAAACTGGAACACTAAACATAGC | |
| **R1_1_2_D1** | CACCAGCTCCTTTATTACCG | |
| R1_2_D2 | GTGGTGTTACCCGTGAACTC | |
| R1_2_R1 | CGTCGCCTAAGTCAAATGAC | |
| **R1_1_2_R2** | TTGGCACTGATCCTTATGAAG | |
| **R1_1_2_CP1** | TTTAGATCGGCGCCGTAACTATGGGCAAGGTTCTTCTTCGTAAGA | |
| R1_2_CP2 | AAGAAAACTGGAACACTAAACATAGCCCCCTCCGTTAAGCTCACGC | |
| **Region 1 ORF1ab primer set 3** | | |
| R1_3_F1 | TTATGGTTGAGCTGGTAGC | This study |
| **R1_3_4_F2** | TGTTATCGACATAGCGAGTGTA | |
| R1_3_C1 | GCTCCTTTATTACCGTTCTTACGAAG | |
| R1_3_C2 | TGGCACTGATCCTTATGAAGATTTTC | |
| R1_3_D1 | ACTGGTATTTCGCCCACATGA | |
| R1_3_D2 | CTGGAACACTAAACATAGCAGTG | |
| R1_3_R1 | ATCGGCGCCGTAACTATGGCCA | |
| R1_3_R2 | TCATTTGACTTAGGCGACGA | |
| R1_3_CP1 | GCTCCTTTATTACCGTTCTTACGAAGAGAACTCGAAGGCATTCAGTA | |
| R1_3_CP2 | TGGCACTGATCCTTATGAAGATTTTCCCTCCGTTAAGCTCACGCATGA | |
| **Region 1 ORF1ab primer set 4** | | |
| R1_4_F1 | AGAACTCGAAGGCATTCAGTA | This study |
| **R1_3_4_F2** | TGTTATCGACATAGCGAGTGTA | |
| R1_4_C1 | TTAGATCGGCGCCGTAACTATGG | |
| R1_4_C2 | CAAGAAAACTGGAACACTAAACATAG | |
| R1_4_D1 | CTTACGAAGAAGAACCTTGC | |
| R1_4_D2 | CAGTGGTGTTACCCGTGAACT | |
| **R1_1_4_R1** | TCGTCGCCTAAGTCAAATGAC | |
| R1_4_R2 | TTGGCACTGATCCTTATGAAGAT | |
| R1_4_CP1 | TTAGATCGGCGCCGTAACTATGGCTCATGTGGGCGAAATACCAGT | |
| R1_4_CP2 | CAAGAAAACTGGAACACTAAACATAGCCTCCGTTAAGCTCACGCATG | |
| **Region 2 ORF1ab primer set 1** | | |
| **R2_1_2_F1*** | CCACAGTACGTCTACAAGC | This study |
| **R2_1_2_F2** | TATGTGGCAACGGCAGTAC | |
| R2_1_C1 | GTAAGCTTTAGCAGCATCTACAGC | |
| **R2_1_3_4_C2** | TGGTCAGGCAATAACAGTTACACC | |
| **R2_1_2_D1** | GCACAGAAAGATAATACAGTTG | |
| **R2_1_2_D2** | AAGCCAATATGGATCAAGAATC | |
| R2_1_R1* | AGTGATTGGTTGTCCCCCAC | |
| R2_1_R2 | AGATGTTGTGTACACACACTG | |
| R2_1_CP1 | GTAAGCTTTAGCAGCATCTACAGCTAATGCAACAGAAGTGCCTGC | |
| R2_1_CP2 | TGGTCAGGCAATAACAGTTACACCAGACAACACGATGCACCACC | |
| **Region 2 ORF1ab primer set 2** | | |
| **R2_1_2_F1** | CCACAGTACGTCTACAAGC | |
| **R2_1_2_F2** | TATGTGGCAACGGCAGTAC | |
| R2_2_C1 | CTTTGTAAGCTTTAGCAGCATCTAC | |

| | | |
|---|---|---|
| R2_2_C2 | GGTCAGGCAATAACAGTTACACCG | |
| **R2_1_2_D1** | GCACAGAAAGATAATACAGTTG | This study |
| **R2_1_2_D2** | AAGCCAATATGGATCAAGAATC | |
| R2_2_R1 | CAATTAGTGATTGGTTGTCCC | |
| R2_2_R2 | GATGTTGTGTACACACACTGG | |
| R2_2_CP1 | CTTTGTAAGCTTTAGCAGCATCTACGTAATGCAACAGAAGTGCCTG | |
| R2_2_CP2 | GGTCAGGCAATAACAGTTACACCGAGACAACACGATGCACCACC | |
| **Region 2 ORF1ab primer set 3** | | |
| R2_3_F1 | AGTTTAGCTGCCACAGTACGTC | |
| **R2_3_4_F2** | CTATGTGGCAACGGCAGTACA | |
| R2_3_C1 | GTAAGCTTTAGCAGCATCTACAGCAA | |
| **R2_1_3_4_C2** | TGGTCAGGCAATAACAGTTACACC | |
| R2_3_D1 | GCACAGAAAGATAATACAGTTGAAT | This study |
| **R2_3_4_D2** | GGAAGCCAATATGGATCAAGAA | |
| **R2_3_4_R1** | ACACAATTAGTGATTGGTTGTCC | |
| R2_3_R2 | AGATGTTGTGTACACACACTGGT | |
| R2_3_CP1 | GTAAGCTTTAGCAGCATCTACAGCAAGGTAATGCAACAGAAGTGCCTGC | |
| **R2_3_4_CP2** | TGGTCAGGCAATAACAGTTACACCGACAACACGATGCACCACCAA | |
| **Region 2 ORF1ab primer set 4** | | |
| R2_4_F1 | CCTAAATAGAGGTATGGTACTT | |
| **R2_3_4_F2** | CTATGTGGCAACGGCAGTACA | |
| R2_4_C1 | TGTAAGCTTTAGCAGCATCTACAGCA | |
| **R2_1_3_4_C2** | TGGTCAGGCAATAACAGTTACACC | |
| R2_4_D1 | GCAGGCACTTCTGTTGCATTACC | This study |
| **R2_3_4_D2** | GGAAGCCAATATGGATCAAGAA | |
| **R2_3_4_R1** | ACACAATTAGTGATTGGTTGTCC | |
| R2_4_R2 | TAAGATGTTGTGTACACACACTG | |
| R2_4_CP1 | TGTAAGCTTTAGCAGCATCTACAGCAAGTTTAGCTGCCACAGTACGTCT | |
| **R2_3_4_CP2** | TGGTCAGGCAATAACAGTTACACCGACAACACGATGCACCACCAA | |
| **Region 3 N gene primer set 1** | | |
| R3_1_F1* | AGAATGGAGAACGCAGTGG | |
| R3_1_F2 | AAATACCATCTTGGACTGAG | |
| R3_1_C1 | TGCCATGTTGAGTGAGAGCGGTG | |
| R3_1_C2 | TAGCAGTCCAGATGACCAAATTGG | |
| R3_1_D1 | AACCAAGACGCAGTATTATTG | This study |
| R3_1_D2 | CTACCGAAGAGCTACCAGAC | |
| R3_1_R1* | CCTCGAGGGAATTTAAGGTC | |
| R3_1_R2 | AAGGCGTTCCAATTAACACC | |
| R3_1_CP1 | TGCCATGTTGAGTGAGAGCGGTGAAACAACGTCGGCCCCAAGG | |
| R3_1_CP2 | TAGCAGTCCAGATGACCAAATTGGTACCGTCACCACCACGAATTC | |
| **Region 3 N gene primer set 2** | | |
| R3_2_F1 | AAACAACGTCGGCCCCAAG | |
| R3_2_F2 | TGGCCCAGTTCCTAGGTAG | |
| R3_2_C1 | TTGTCCTCGAGGGAATTTAAGGTC | |
| **R3_2_3_C2** | ACCGAAGAGCTACCAGACGAATTC | |
| R3_2_D1 | TTGCCATGTTGAGTGAGAGC | This study |
| R3_2_D2 | GTGGTGGTGACGGTAAAATG | |
| R3_2_R1 | CTATTGGTGTTAATTGGAACG | |
| R3_2_R2 | CAGATGACCAAATTGGCTAC | |
| R3_2_CP1 | TTGTCCTCGAGGGAATTTAAGGTCAATACTGCGTCTTGGTTCACC | |
| R3_2_CP2 | ACCGAAGAGCTACCAGACGAATTCAGAAATACCATCTTGGACTGAG | |
| **Region 3 N gene primer set 3** | | |
| R3_3_F1 | CGATCAAAACAACGTCGGCC | |
| R3_3_F2 | TCTGGCCCAGTTCCTAGGTA | |
| R3_3_C1 | TTGTCCTCGAGGGAATTTAAGGTCTT | |
| **R3_2_3_C2** | ACCGAAGAGCTACCAGACGAATTC | |
| R3_3_D1 | CTTGCCATGTTGAGTGAGAGC | This study |
| R3_3_D2 | GTGGTGGTGACGGTAAAATGAA | |
| R3_3_R1 | CTATTGGTGTTAATTGGAACGCC | |
| R3_3_R2 | TCCAGATGACCAAATTGGCTAC | |
| R3_3_CP1 | TTGTCCTCGAGGGAATTTAAGGTCTTCAATAATACTGCGTCTTGGTTCAC | |
| R3_3_CP2 | ACCGAAGAGCTACCAGACGAATTCTAGAAATACCATCTTGGACTGAGA | |
| **Region 3 N gene primer set 4** | | |
| R3_4_F1 | GCAGTAACCAGAATGGAGAAC | |
| R3_4_F2 | TTCTGGCCCAGTTCCTAGGTA | |
| R3_4_C1 | CTTGCCATGTTGAGTGAGAGCGGT | |
| R3_4_C2 | CAAATTGGCTACTACCGAAGAGCTAC | |
| R3_4_D1 | GAACCAAGACGCAGTATTATTG | This study |
| R3_4_D2 | AATTCGTGGTGGTGACGGTAA | |
| R3_4_R1 | TCCTCGAGGGAATTTAAGGTCT | |
| R3_4_R2 | AAGGCGTTCCAATTAACACCAATA | |

| | |
|---|---|
| R3_4_CP1 | CTTGCCATGTTGAGTGAGAGCGGTCAAAACAACGTCGGCCCCAAG |
| R3_4_CP2 | CAAATTGGCTACTACCGAAGAGCTACTAGAAATACCATCTTGGACTGAGAT |

| **LAMP primer name** | **Sequence 5' --> 3'** | |
|---|---|---|
| **Region 1 ORF1ab primer set** | | |
| ORF1a-A-F3 | CTGCACCTCATGGTCATGTT | Zhang et al. 2020 Rapid Molecular Detection of SARS-CoV-2 (COVID-19) Virus RNA Using Colorimetric LAMP |
| ORF1a-A-B3 | AGCTCGTCGCCTAAGTCAA | |
| ORF1a-A-FIP | GAGGGACAAGGACACCAAGTGTATGGTTGAGCTGGTAGCAGA | |
| ORF1a-A-BIP | CCAGTGGCTTACCGCAAGGTTTTAGATCGGCGCCGTAAC | |
| ORF1a-A-LF | CCGTACTGAATGCCTTCGAGT | |
| ORF1a-A-LB | TTCGTAAGAACGGTAATAAAGGAGC | |
| **Region 3 N gene primer set** | | |
| GeneN-A-F3 | TGGCTACTACCGAAGAGCT | Zhang et al. 2020 Rapid Molecular Detection of SARS-CoV-2 (COVID-19) Virus RNA Using Colorimetric LAMP |
| GeneN-A-B3 | TGCAGCATTGTTAGCAGGAT | |
| GeneN-A-FIP | TCTGGCCCAGTTCCTAGGTAGTCCAGACGAATTCGTGGTGG | |
| GeneN-A-BIP | AGACGGCATCATATGGGTTGCACGGGTGCCAATGTGATCT | |
| GeneN-A-LF | GGACTGAGATCTTTCATTTTACCGT | |
| GeneN-A-LB | ACTGAGGGAGCCTTGAATACA | |

| **rt-PCR primer name** | **Sequence 5' --> 3'** | |
|---|---|---|
| **Region 3 N gene primer set** | | |
| WH-NIC N-F | CGTTTGGTGGACCCTCAGAT | WHO Molecular assays to diagnose COVID-19: Summary table of available protocols- National Institute of Health, Thailand |
| WH-NIC N-R | CCCCACTGCGTTCTCCATT | |
| WH-NIC N-P | FAM-CAACTGGCAGTAACCA- BHQ1 | |

**Supplementary Table 2:** List of synthesised gene fragments used as DNA/RNA template for MCDA, LAMP and rt-PCR. Blue indicates universal M13 adapters while red depicts the sequence for T7 promoter.

| Gene fragment name | Position in NC_045512.2 | Gene target | Sequence 5' --> 3 ' |
|---|---|---|---|
| Region 1 | 416-931 | ORF1ab | TGTAAAACGACGGCCAGTTAATACGACTCACTATAGTGTGGCTTAGTAGAAGTTGAAAAAGGCGTTTTGCCTCAACTTGAACAGCCCTATGTGTTCATCAAACGTTCGGATGCTCGAACTGCACCTCATGGTCATGTTATGGTTGAGCTGGTAGCAGAACTCGAAGGCATTCAGTACGGTCGTAGTGGTGAGACACTTGGTGTCCTTGTCCCTCATGTGGGCGAAATACCAGTGGCTTACCGCAAGGTTCTTCTTCGTAAGAACGGTAATAAAGGAGCTGGTGGCCATAGTTACGGCGCCGATCTAAAGTCATTTGACTTAGGCGACGAGCTTGGCACTGATCCTTATGAAGATTTTCAAGAAAACTGGAACACTAAACATAGCAGTGGTGTTACCCGTGAACTCATGCGTGAGCTTAACGGAGGGGCATACACTCGCTATGTCGATAACAACTTCTGTGGCCCTGATGGCTACCCTCTTGAGTGCATTAAAGACCTTCTAGCACGTGCTGGTAAAGCTTCATGCACTTTGTCCGAACAACTGGACTTTATTGGTCATAGCTGTTTCCTG |
| Region 2 | 12869-13388 | ORF1ab | TGTAAAACGACGGCCAGTTAATACGACTCACTATAGACTGGTACTATCTATACAGAACTGGAACCACCTTGTAGGTTTGTTACAGACACACCTAAAGGTCCTAAAGTGAAGTATTTATACTTTATTAAAGGATTAAACAACCTAAATAGAGGTATGGTACTTGGTAGTTTAGCTGCCACAGTACGTCTACAAGCTGGTAATGCAACAGAAGTGCCTGCCAATTCAACTGTATTATCTTTCTGTGCTTTTGCTGTAGATGCTGCTAAAGCTTACAAAGATTATCTAGCTAGTGGGGGACAACCAATCACTAATTGTGTTAAGATGTTGTGTACACACACTGGTACTGGTCAGGCAATAACAGTTACACCGGAAGCCAATATGGATCAAGAATCCTTTGGTGGTGCATCGTGTTGTCTGTACTGCCGTTGCCACATAGATCATCCAAATCCTAAAGGATTTTGTGACTTAAAAGGTAAGTATGTACAAATACCTACAACTTGTGCTAATGACCCTGTGGGTTTTACACTTAAAAACACAGTCTGTACCGTCTGCGGTAGGTCATAGCTGTTTCCTG |
| Region 3 | 28246-28747 | N | TGTAAAACGACGGCCAGTTAATACGACTCACTATAGTAGATTTCATCTAAACGAACAAACTAAAATGTCTGATAATGGACCCCAAAATCAGCGAAATGCACCCCGCATTACGTTTGGTGGACCCTCAGATTCAACTGGCAGTAACCAGAATGGAGAACGCAGTGGGGCGCGATCAAAACAACGTCGGCCCCAAGGTTTACCCAATAATACTGCGTCTTGGTTCACCGCTCTCACTCAACATGGCAAGGAAGACCTTAAATTCCCTCGAGGACAAGGCGTTCCAATTAACACCAATAGCAGTCCAGATGACCAAATTGGCTACTACCGAAGAGCTACCAGACGAATTCGTGGTGGTGACGGTAAAATGAAAGATCTCAGTCCAAGATGGTATTTCTACTACCTAGGAACTGGGCCAGAAGCTGGACTTCCCTATGGTGCTAACAAAGACGGCATCATATGGGTTGCAACTGAGGGAGCCTTGAATACACCAAAAGATCACATTGGCACCCGCAATCCTGCTAACAATGCTGCAATCGTGGGTCATAGCTGTTTCCTG |

BLUE = universal M13
RED = T7 promoter
